# Association of physical activity and the risk of COVID-19 hospitalization: a dose–response meta-analysis

**DOI:** 10.1101/2022.06.22.22276789

**Authors:** Dan Li, Shengzhen Jin, Songtao Lu

**Affiliations:** School of Sports, Wuhan University of Science and Technology, Wuhan 430081, China; Tennis College, Wuhan sports university, Wuhan 430079, China; School of Physical Education and Sports, Central China Normal University, Wuhan 430079, China

**Author notes:** These authors contributed equally to this work (Dan Li, Shengzhen Jin).

## Abstract

**Background:** Many people have experienced a high burden due to the spread of the coronavirus disease (COVID-19) and its serious consequences for health and everyday life. Prior studies have reported that physical activity (PA) may lower the risk of COVID-19 hospitalization. The present meta-analysis (PROSPERO registration number: CRD42022339672) explored the dose– response relationship between PA and the risk of COVID-19 hospitalization.

**Methods:** Epidemiological observational studies on the relationship between PA and the risk of COVID-19 hospitalization were included. Categorical dose–response relationships between PA and the risk of COVID-19 hospitalization were assessed using random effect models. Robust error meta-regression models assessed the continuous relationship between PA (metabolic equivalent [MET]–h/week) and COVID-19 hospitalization risk across studies reporting quantitative PA estimates.

**Results:** Seventeen observational studies (cohort\case–control\cross-section) met the criteria for inclusion in the meta-analysis. Categorical dose-relationship analysis showed a 40% (risk ratio (RR) 0.60, 95% confidence intervals (CI): 0.48–0.71) reduction in the risk of COVID-19 hospitalization compared to the lowest dose of PA. The results of the continuous dose–response relationship showed a non-linear inverse relationship (Pnon-linearity < 0.05) between PA and the risk of COVID-19 hospitalization. When total PA was less than or greater than 10 Met-h/week, an increase of 4 Met-h/week was associated with a 14% (RR = 0.83, 95%CI: 0.85–0.87) and 11% (RR = 0.89, 95%CI: 0.87–0.90) reduction in the risk of COVID-19 hospitalization, respectively.

**Conclusions:** There was an inverse non-linear dose–response relationship between PA level and the risk of COVID-19 hospitalization. Doses of the guideline-recommended minimum PA levels by WTO may be required for more substantial reductions in the COVID-19 hospitalization risk.

## 1. Introduction

The coronavirus disease 2019 (COVID-19) outbreak continues worldwide. As of 7 May 2022, COVID-19 has caused 51,587,3758 infections and 6,272,357 deaths worldwide [1]. It is essential to identify high-risk groups that require special attention under these conditions [2]. For non-communicable disease outcomes, lifestyle risk factors have been consistently associated with morbidity, mortality, and loss of disease-free life [3,4]. For example, physical inactivity and smoking appear to be independently associated with a higher risk of community-acquired pneumonia morbidity and mortality [5,6].

It is also well established that the risk of developing respiratory disease is much higher in people with low physical activity (PA), whereas COVID-19 patients with a physically inactive lifestyle (e.g., sedentary behavior) are more likely to be hospitalized and have a greater likelihood of poor clinical outcomes [7]. Moreover, it has previously been shown that regular physical activity and higher physical fitness levels enhance immune function and, therefore, might reduce susceptibility to COVID-19 infection and infection severity [8,9]. Recent studies retrospectively evaluating cohorts of COVID-19 positive adults have described the benefit of regular physical activity in decreasing the incidence of adverse outcomes in confirmed cases of COVID-19 [10,11,12].

However, research on such topics is just emerging, and the impact of PA on the infectious and clinical outcomes of COVID-19 remains unclear. The protective effects of different levels of physical activity against COVID-19 are controversial. Rahmati et al. [13] conducted a meta-analysis on this topic, which did not address the controversy regarding the protective effects of different levels of physical activity. In addition, Rahmati et al. [13] classified the case–control group as a cross-sectional study. In a metaanalysis, they assumed cardiopulmonary function as physical activity, which inevitably led to unconvincing results. Finally, the study only used the binary variables of physical activity included in the literature to analyze the outcome variables, ignoring the moderate dose in the multi-level doses, making it difficult to explain the heterogeneity generated by the meta-analysis studies.

Furthermore, no systematic review or meta-analysis has reported the exact dose–response relationship between pre-diagnosis PA and COVID-19 hospitalization. Consequently, there is still substantial uncertainty regarding the association between prediagnosis PA levels and hospitalization due to COVID-19 among the general population. To precisely quantify the association between pre-diagnosis PA and the risk of COVID-19 hospitalization, we conducted a systematic review and meta-analysis of observational studies published up to May 8, 2022.

## 2. Materials and Methods

### 2.1. Inclusion Criteria

The criteria for inclusion, and each article determined for inclusion, were discussed by three authors, and the discussions on inclusion and exclusion occurred more than three times. The inclusion criteria were as follows: (1) studies published as epidemiological observational cohort studies, case controls, and cross-sectional design investigation studies; (2) studies providing at least an odds ratio, relative risk (RR or HR), and 95% confidence interval (95%CI) between the level of physical activity and the risk of hospitalization for COVID-19, or raw data provided to calculate these indicators. The repeated literature was excluded. Only the latest studies were selected if they were conducted at different time points in the same cohort. Additionally, if multiple articles were published in the same group, we chose articles where subjects were followed for a longer time or with a larger sample size.

### 2.2. Search Strategy

We searched PubMed (1980 to the present) and the Web of Science database (1980 to the present) for literature on the relationship between physical activity and the risk of COVID-19 hospitalization. The search strategy used keywords such as “exercise or physical activity or sport or walking or motor activity”, “COVID-19 or SARS-CoV-2”, and “severe or hospitalization”. These searches were screened for cohort studies, case controls, and cross-sectional design studies. The latest search date was April 2022, and there was no language limit. The reference lists of the selected and related review articles were screened to identify potentially relevant studies. All searches were conducted independently by two authors, and the differences were resolved by group discussion.

### 2.3. Quality Assessment

The Newcastle–Ottawa Scale (NOS) was used to evaluate literature quality, and scores of 0–3, 4–6, and 7–9 were determined as low, moderate, and high quality, respectively [14]. Each article was evaluated independently by two authors and cross-checked. In the group meeting, the results were publicized, and the reasons for the score of each item were specified. If the evaluation of the literature quality was inconsistent, the group focused on solving the final score of its quality.

### 2.4. Synthesis Methods

Stata16.0 software was used for the meta-analysis. The p-value was set at p < 0.05, and all tests were double-sided. The effect value-adjusted risk ratio (RR) and 95% confidence interval (CI) of the group with the highest dose of PA (physical activity) compared to the control group with the lowest dose of PA in each study were combined. The combined effect values were calculated using a random-effects model. Heterogeneity was assessed and described using I2 statistics as the percentage of variation in the study; I2 values of 25%, 50%, and 75% represented low, moderate, and high levels of heterogeneity, respectively [15]. Egger and Begg tests were used to determine publication bias. During the sensitivity analysis, each study was deleted one by one to check whether the combined effect of the remaining studies had changed [15]. The subgroup meta-analysis was conducted according to PA intensity classification (LPA-light intensity physical activity, VPA-vigorous-intensity physical activity, MVPA-moderate-to-vigorous physical activity, MPA-moderate-intensity physical activity), sex, age, study area, study quality, and adjustment for confounding factors. Meta-regression was used to examine the heterogeneity among studies.

According to categorical and continuous dose PA, this study analyzed the dose–response relationship with the risk of COVID-19 hospitalization. Categorical doses were divided into dichotomous and multi-classified doses shown in the study, and the combined effect value RR was generated by comparing the highest and lowest doses. To analyze the continuous dose–response relationship, we calculated the total weekly dose of PA for each effect value RR based on the PA intensity, duration, and weekly frequency of the baseline survey provided in the literature. Furthermore, we assumed that the dose remained at this level during the followup survey. To determine the exposure value of the included dose, the median was set as the determined dose. If the development interval was < 0.5, it was set to 0.25. If the upper open interval was ≥ 1, the difference between the intermediate dose intervals was 0.25, and the exposure value was set to 1.25 [16]. Met-h/week was considered as the final unit of analysis. These are combined absolute indices of intensity, duration, and frequency used to calculate exposures to Met units not directly reported in the literature. Met, a physiological measure of PA energy, is defined as energy expenditure per kilogram of body weight per hour: 1 Met = 1 kcal/kg∗h. To address the differences in PA units in different studies, we used Ainsworth et al. ‘s classification [16], classifying PA into low-intensity LPA (3 Mets, such as walking exercise), moderate-intensity MPA(4 Mets), and high-intensity VPA (8 Mets). We then converted the duration of a particular PA intensity (h/week) to Met-h/week in combination with the frequency of the week [17].

To establish the dose–response relationship between PA and the risk of hospitalization for COVID-19, robust error metaregression (REMR) was used for model fitting [18]. The REMR approach is based on a “one-stage” framework that treats each study as a cluster and fits the revised regression to the average PA dose across the entire dataset. In addition, the method also uses the inverse variance method to weight each dose-specific effect in the data and balances heteroscedasticity in the REMR model to ensure unbiased estimation of parameters. Finally, we used restricted cubic splines as connection functions to fit the linear and non-linear dose–response models. Based on the dose-centralization treatment, the independent variable PA dose of the model was set as three nodes (0, 6.75, and 21), including two regression splines. The χ2 test was used to test the hypothesis that the regression coefficient of the second regression spline is significant (p < 0.05), indicated by a linear or non-linear dose– response relationship. A dose–response relationship curve was drawn using the Stata software XBLC command [16].

## 3. Results

### 3.1. Study Selection and Characteristics

In total, 170 articles were preliminarily identified. According to the literature inclusion and exclusion criteria formulated in this study, 17 studies, including seven cohort studies, five case–control studies, and five cross-sectional design studies, were finally included. There were 1,038,768 subjects and about 3022 hospitalized COVID-19 cases (some studies had not reported the number of cases). The steps for retrieval and inclusion are shown in Fig 1. The characteristics of the literature are listed in Table 1. Among the 17 studies, 3 were from North America [10,19,20], 7 were from Asia [11,12,21,22,23,24,25], 6 were from Europe [26,27,28,29,30,31], and 1 was from Oceania [32]. The NOS was used to score the included studies. Eight studies were considered high quality, with a score greater than or equal to seven, and the other nine were considered medium quality.

**Table 1.** Summary of the extracted studies.

| Author (year) | Country | Study type | Case\total | Age (SD) | female % | Measurement and Categories of PA | adjustment * | QA |
| --- | --- | --- | --- | --- | --- | --- | --- | --- |
| AlKetbi (2021) | AUH | Cohort | 135\641 | 44(NP) | 36% | Self-report PA: five Categories(times\w) | NP* | 5 |
| Brandenburg (2021) | CA | Case-control | NP\263 | 86%<65 | 57% | PA-R: No, Moderate, <1 h of vigorous, >1 h of vigorous | 2 5 6 8 9 12 15 | 7 |
| Bielik(2021) | SLK | Cohort | 104\2343 | 18 to 65 | 49% | physically active($\geq 3$ times\w), cold-water swim | NP | 5 |
| Ekblom-Bak(2021) | SWE | Case-control | 172\407131 | 49.9(NP) | 30% | Never/irregular, 1–2 times/w, $\geq 3$ times/w | 1 2 5 16 | 7 |
| Katsoulis (2021) | UK | Cohort | NP\ 85308 | 18-69(NP) | NP | Self-report PA: Low, Gentle, Moderate, Vigorous | 1 2 7 9 | 7 |
| Hamer(2020) | UK | Cohort | 760\387109 | 56.2± 8.0 | 55.1% | IPAQ: Sufficient\ Insufficient\ None | 1 2 6 8 9 | 7 |
| Halabchi (2021) | Iran. | Cross-Section | 60\4694 | 36.45±9.77 | 55% | Regular sports participation (yes/no) | NP | 6 |
| Hamdan(2021) | PLE | Cross-Section | 59\300 | 30.5±12.2 | 55.0% | PA questionnaire: yes\no | NP | 5 |
| Lee (2021) | KR | Cohort | 277\118768 | NP | 51.2% | Self-report PA: None, Gentle, Moderate, Vigorous | 1 2 5-9 14 16 |  |
| Latorre-Roman(2021) | Spanish | Cross-Section | NP\420 | 33 (20–54) | 52.6% | IPAQ: Moderate PA>150 min\w, 30–150 min\w, None | 1 | 6 |
| Malisoux(2022) | LUX | Cohort | 106\452 | 42 (31–51) | 48% | PA questionnaire: yes\no | 1 2 3 5 16 18 19 | 7 |
| Maltagliati(2021) | France | Case-control | 66\3139 | 69.3 ± 8.5 | 53% | four-point PA scale ranging (>1, 1; <1; 0\week) | 1 2 5 7 8 12 19 | 9 |
| Souza et al(2021) | Brazil | Cross-Section | 91\938 | NP(NP) | 33.4% | IPAQ: Sufficient>150 min/w(moderate), Insufficient | 1 2 19 | 7 |
| Sallis (2021) | USA | Cohort | 1199\2970 | 47±16.97 | 61.9% | UPAG: active, inactive, some activity | 1-15 | 9 |
| Steenkamp (2021) | ZA | Cohort | NP\65361 | 41±12.1 | 48.2% | Low activity, Moderate activity, High activity | 1 2 6 8 9 14 15 | 8 |
| Tavakol(2021) | Iran | Cross-Section | 64\188 | 18-75(NP) | 52.7% | GPAQ: Low, Moderate to high | NP | 6 |
| Yuan(2021) | CN | Case-control | 29\164 | 61.8 ± 13.6 | 48.8% | Self-report PA: Inactivity, activity | NP | 6 |

Case \ total: Number of cases and total sample size; Age characteristics: Single value indicates mean age, others are age range.\* Adjustment factors: 1 age, 2 gender, 3 socioeconomic status, 4 race, 5BMI, 6 cardiovascular disease, 7 cancer, 8 diabetes, 9 hypertension, 10 use of antihypertensive drugs, 11 corticosteroids, 12 chronic lung / respiratory disease, 13 liver disease, 14 HIV, 15 end-stage renal disease and immune disease, 16 smoking, 17 alcohol,18 sedentary behaviour , 19 comorbidities.NP \*: Not reported. AUH: AUH , ZA: South Africa, UK: UK, AUH: UAE, CA: Canada, SWE: Switzerland, Iran: Iran, KR: Korea, ESP: Spain, USA: US, ZA: New Zealand, CN: China, SLK: Slovak, LUX: Luxembourg, PLE: Palestine. IPAQ: International Physical Activity Questionnaire. PA-R: physical activity rating questionnaire. UPAG: US Physical Activity Guidelines. GPAQ: Global physical activity questionnaire.

**Fig 1.**
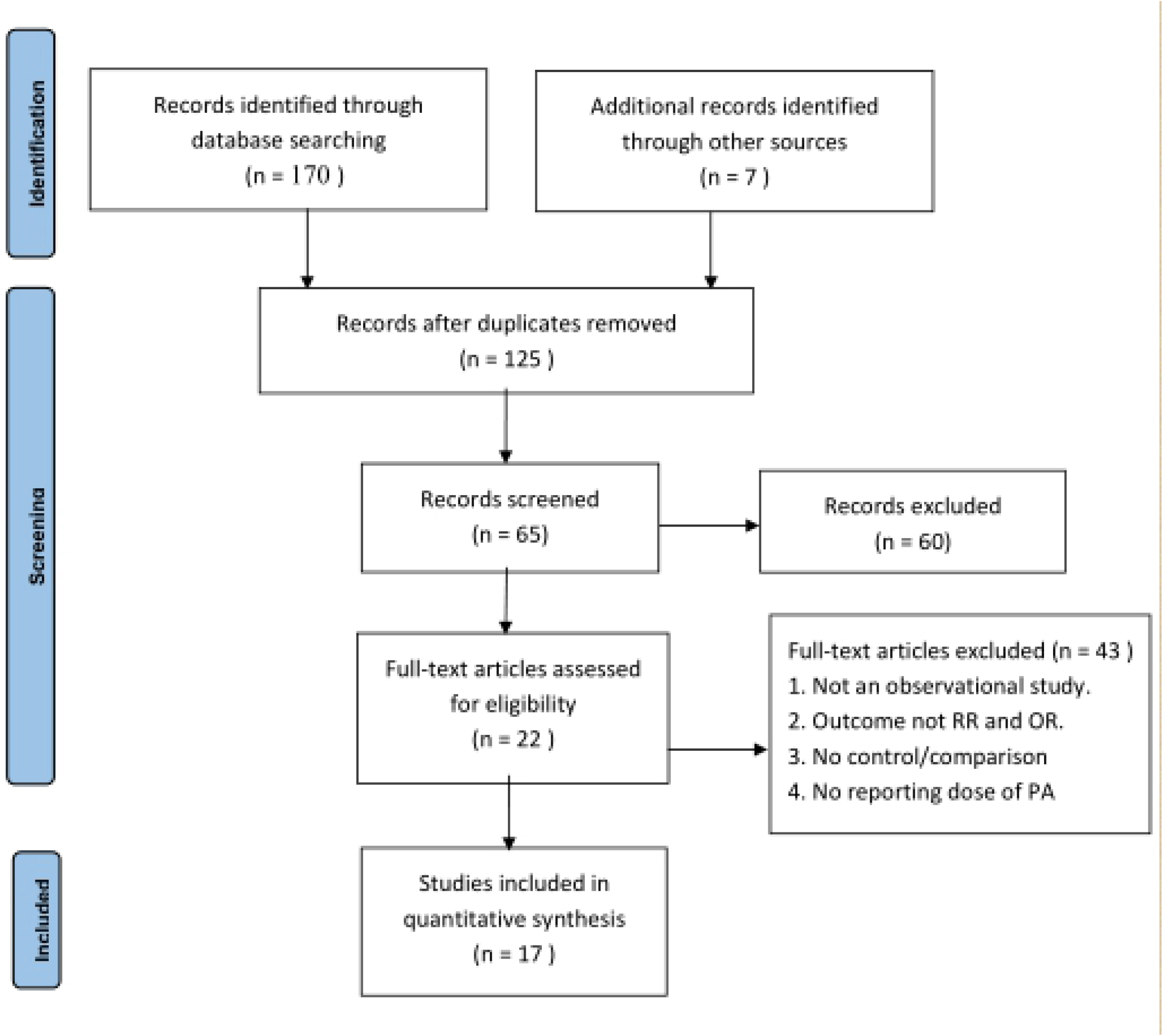
Flow diagram of studies considered for inclusion in the systematic review.

### 3.2. Categorical analysis between PA and COVID-19 hospitalization

Compared with the lowest PA dose, the highest PA in the included studies reduced the risk of COVID-19 hospitalization by 40% (RR = 0.60, 95%CI: 0.48, 0.71). The heterogeneity test result was I2 = 66.22% (p < 0.01), indicating significant heterogeneity of the study results (Fig 2). The pooled effect size results did not change significantly after excluding each study from the sensitivity analysis (Supporting information S1 Fig). Published bias analysis with Begg’s test p = 0.54 > 0.05, Egger test p = 1.330 > 0.05, and funnel diagram also showed no significant published bias (Supporting information S2 Fig). The effect values of the cohort study, case–control study, and cross-sectional design study were 0.63 (95%CI:0.54,0.71), 0.59 (95%CI: 026, 0.91), and 0.58 (95%CI: 0.42, 0.74), respectively.

**Fig 2.**
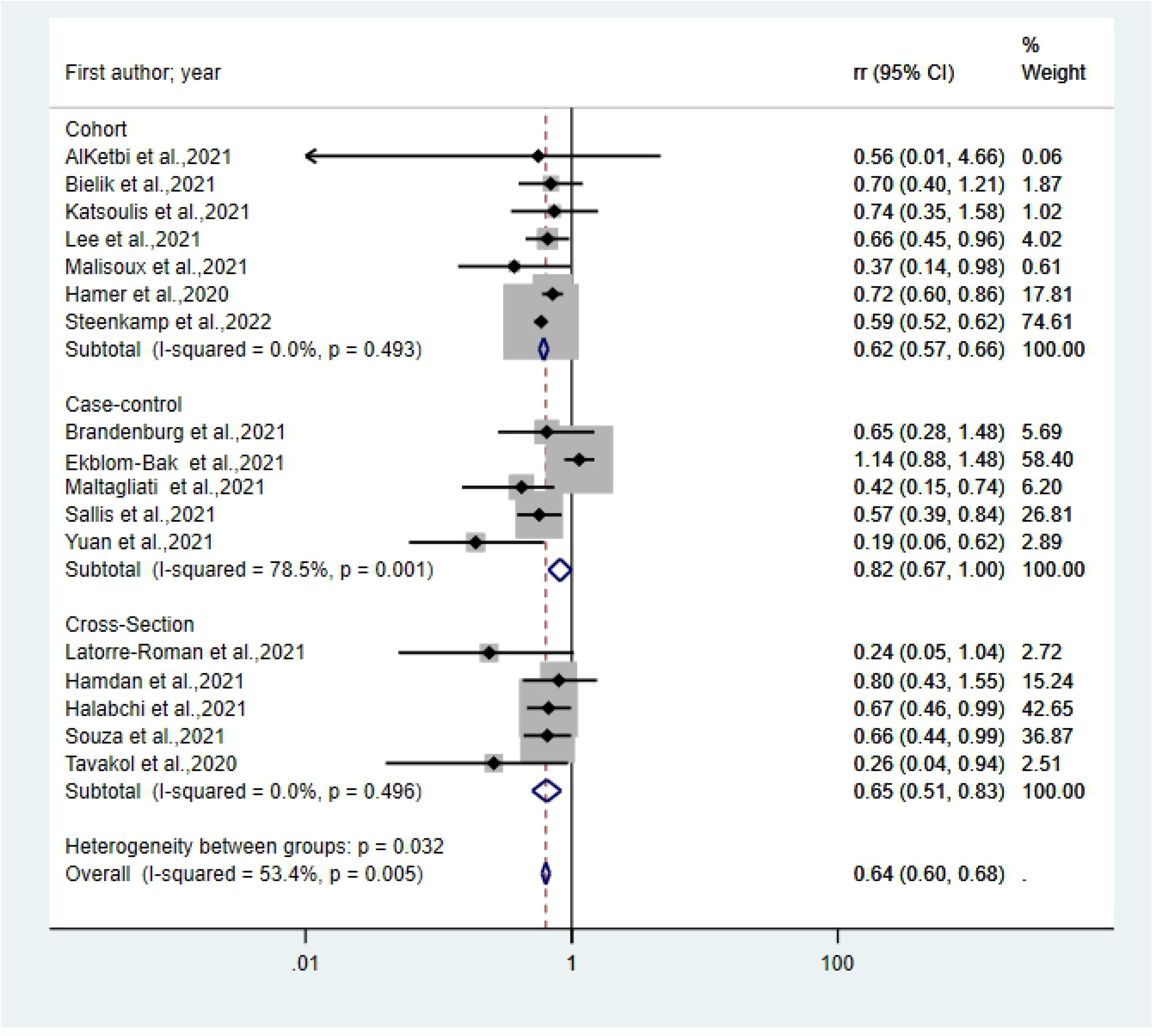
Forest plot showing categorical analysis between PA and COVID-19 hospitalization.

As for the source of heterogeneity (presented in Table 2), between-group heterogeneity only appeared in the comparative analysis of the relationship between PA at different dose levels and the risk of hospitalization for COVID-19 (Pb < 0.01), indicating that PA at different doses significantly reduces the risk of hospitalization. Within-group heterogeneity appeared in the multi-dose PA, case–control study, high quality, European, adjustment for age, sex, adjustment for high blood pressure, adjustment for diabetes, adjustment for cancer, and cardiovascular diseases subgroup, illustrating that the subgroup study effect value of the results may not be stable. The results may be affected by other factors.

**Table 2.** Subgroup analysis

| Subgroup |  | N | RR(95% CI) | I<sup>2</sup> (%) | p<sup>a</sup>* | p<sup>b</sup>* |
| --- | --- | --- | --- | --- | --- | --- |
| Total effect size (highest vs. lowest) |  | 17 | 0.60 (0.48,0.71) | 66.22 | 0.01 |  |
| PA Categories | binary does * | 7 | 0.51(0.33,0.69) | 45.43 | 0.09 | 0.24 |
|  | multi-dose * | 10 | 0.65(0.51,0.79) | 68.34 | 0.02 |  |
| multi-class dose comparison* | highest vs. lowest | 12 | 0.59(0.55,0.63) | 0 | 0.86 | 0.01< |
|  | moderate vs. lowest | 13 | 0.75(0.65,0.85) | 26.88 | 0.33 |  |
| Study type | cohort | 7 | 0.63(0.54,0.71) | 20.83 | 0.53 | 0.85 |
|  | Case-control | 5 | 0.59(0.26,0.91) | 82.07 | 0.00 |  |
|  | Cross-section | 5 | 0.58(0.42,0.74) | 2.05 | 0.28 |  |
| Study quality | ≥ 7 | 8 | 0.67(0.53,0.81) | 70.32 | 0.02 | 0.1 |
|  | 7< | 9 | 0.49(0.32,0.66) | 38.93 | 0.15 |  |
| Different continent | Europe | 6 | 0.68(0.43,0.94) | 68.68 | 0.02 | 0.46 |
|  | Asia | 7 | 0.49(0.30,0.68) | 46.31 | 0.11 |  |
|  | others (America) | 4 | 0.59(0.54,0.64) | 0 | 0.96 |  |
| Adjusted confounding factor |  |  |  |  |  |  |
| age | yes | 10 | 0.63(0.51,0.75) | 66.73 | 0.01 | 0.36 |
|  | no | 7 | 0.51(0.29,0.74) | 43.20 | 0.14 |  |
| sex | yes | 10 | 0.65(0.54,0.78) | 59.93 | 0.03 | 0.15 |
|  | no | 7 | 0.47(0.25,0.69) | 44.89 | 0.12 |  |
| BMI | yes | 7 | 0.66(0.48,0.84) | 67.76 | 0.02 | 0.29 |
|  | no | 10 | 0.54(0.40,0.87) | 43.12 | 0.15 |  |
| Adjusted baseline disease |  |  |  |  |  |  |
| hypertension | yes | 8 | 0.58(0.31,0.85) | 10.28 | 0.57 | 0.83 |
|  | no | 9 | 0.61(0.55,0.67) | 71.22 | 0.00 |  |
| diabetes | yes | 8 | 0.61(0.55,0.87) | 11.67 | 0.49 | 0.91 |
|  | no | 9 | 0.59(0.34,0.84) | 67.09 | 0.00 |  |
| cardiovascular | yes | 8 | 0.61(0.55,0.87) | 11.67 | 0.49 | 0.91 |
|  | no | 9 | 0.59(0.34,0.84) | 67.09 | 0.00 |  |
| cancer | yes | 6 | 0.57(0.45,0.69) | 0 | 0.70 | 0.75 |
|  | no | 11 | 0.61(0.43,0.79) | 80.95 | 0.00 |  |

Multi-dose comparison \*: The highest dose is higher than the 10 h-met / week and the lowest dose, and the moderate dose is lower than 10 h-met / week. The binary dose is generally expressed as exercise and non-exercise; \* multiple dose grouping represents the highest, relative and lowest dose comparison, at least three lever; P<sup>a</sup> and P<sup>b</sup> represent heterogeneity within and between subgroups, respectively.

### 3.3. Continuous dose-response relationship between PA and COVID-19 hospitalization

Fig 3 shows the continuous dose–response relationship. The results showed a negative non-linear relationship between PA and the risk of hospitalization for COVID-19 (Pnon-linearity < 0.01). When PA < 10 Met-h/week, an increase of 4 Met-h/week (1 h of moderate-intensity or 1/2 h of high-intensity) was associated with a 14% reduction in the risk of hospitalization for COVID-19 (p < 0.01, RR = 0. 86, 95%CI: 0.85–0.87). When PA > 10 Met-h/week, the risk of hospitalization for COVID-19 decreased by 11% for each additional 4 Met-h/week (p < 0.01, RR = 0.89, 95%CI: 0.87–0.90).

**Fig 3.**
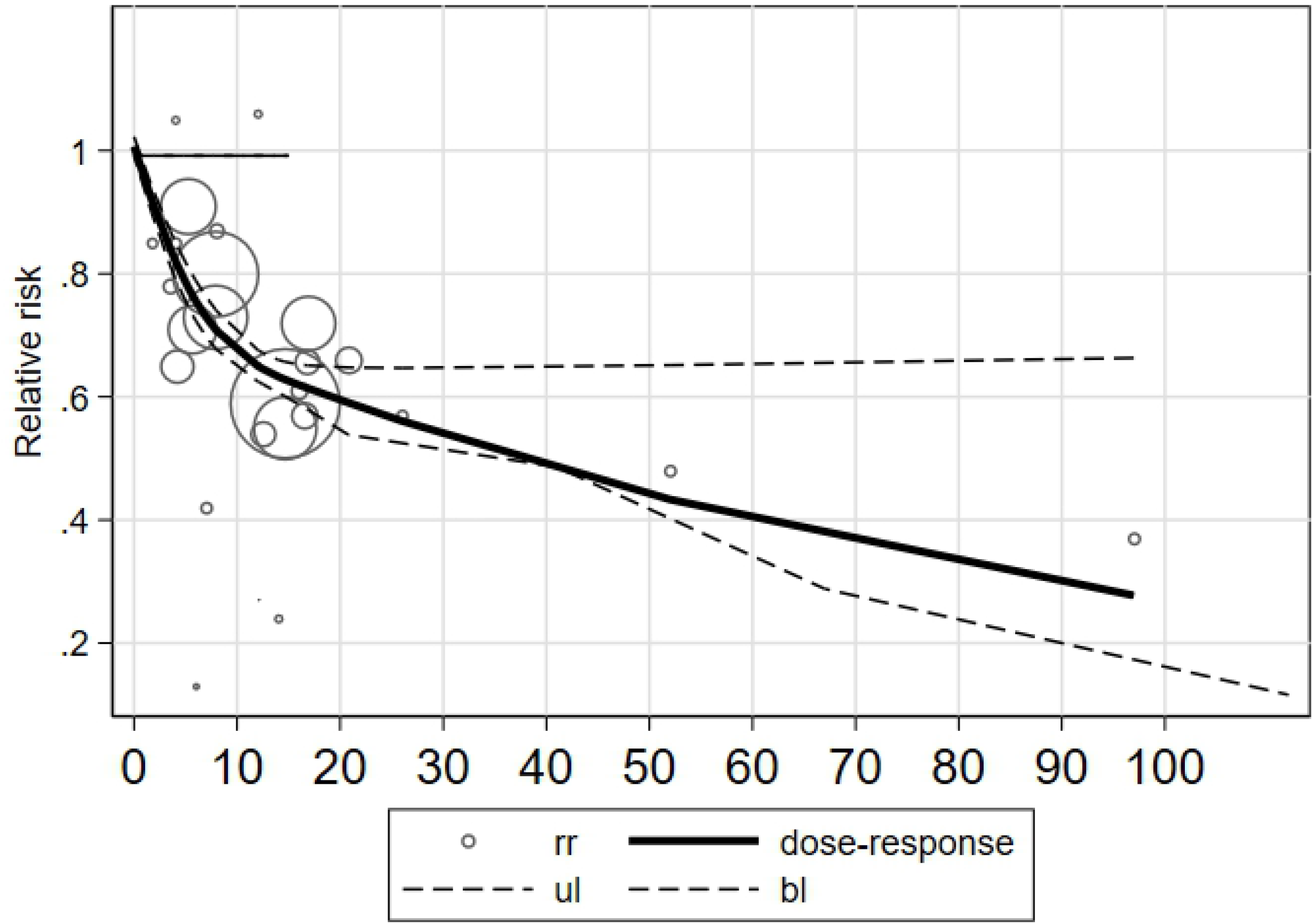
Continuous dose-response relationship between PA and COVID-19 hospitalization.

## 4. Discussion

This study is the first dose–response meta-analysis of the relationship between PA and the risk of COVID-19 hospitalization. The literature included observational studies on the relationship between PA and the risk of COVID-19 hospitalization. Through a metaanalysis of the categorical dose, our main conclusion is that the risk of COVID-19 hospitalization is reduced by 40% compared with the lowest dose of PA. For continuous dose–response analysis, we confirmed that the relationship between PA and the risk of COVID-19 hospitalization is non-linear and inverse. For every four Met-h\week PA increase, the risk of COVID-19 hospitalization decreased by 11–14%. Sensitivity and published bias analyses further support these results, and these main quantitative features have important clinical significance.

The dose–response association between PA and the risk of COVID-19 hospitalization has been previously reported. As for the categorical analysis, Rahmati et al. observed that PA was significantly associated with a reduction in COVID-19 hospitalization compared with control (RR = 0.58) by meta-analysis [13]. In the present study, we observed a similar RR of 0.60. However, we included multiple categorical analysis data with different PA dose levels to analyze the relationship between PA and the risk of COVID-19 hospitalization. We observed significantly different protective effects of varying PA levels on the hospitalization risk of COVID-19 by heterogeneity analysis. Compared to the results obtained by Rahmati et al., our observations are more abundant.

As for the continuous analysis, Malisoux et al. observed an inverse dose–response association between PA and the risk of moderate COVID-19 illness, and increased PA was associated with a slightly lower risk of moderate illness (OR:0.99) [25]. In the present study, we observed a similar inverse dose–response association between PA and the risk of COVID-19 hospitalization. An increase of 4 Met-h/week PA was associated with a 12–17% (RR = 0.83–0.88) reduction in the risk of COVID-19 hospitalization. A 4 Met-h/week PA is equivalent to one hour MPA or half an hour high VPA; our results are more specific and close to a practical exercise. Otherwise, the observed dose–response relationship between PA and the risk of COVID-19 hospitalization was non-linear. We assumed that after 10 Met-h/week, the degree of enhancement in lowering the risk of moderate COVID-19 illness when PA increase is weakened. However, the threshold is 30 Met-h/week according to the j-shaped association of PA and risk of moderate illness by Malisoux et al.. [25]. Meanwhile, our meta-analysis findings are more robust and specific, and we observed increasing PA benefits for the inpatient burden due to COVID-19.

This difference in the magnitude of risk reduction for COVID-19 hospitalization could be related to differences in the mechanism through which PA modifies the risk of respiratory viral infections. This is supported by previous studies that showed a stronger association between maximal fitness exercise capacity and the risk of hospitalization due to COVID-19 [33]. Exercise capacity is an important index for measuring overall health and the ability of the body to cope with external stressors. More specifically, it is an important index to bear the burden on the heart and lungs [33]. However, PA greatly influences exercise capacity; more specifically, regular moderate-intensity to vigorous-intensity aerobic exercise daily can improve exercise capacity. In addition, the beneficial effects of regular PA on the immune system may involve several mechanisms, including enhanced immunosurveillance, reduced systemic inflammation, improved regulation of the immune system, and delayed onset of immunosenescence [34,35]. A recent meta-analysis investigated the effects of regular PA on the immune system [36]. This study showed that moderate to vigorous intensity exercise (e.g., walking, running, cycling) is overall beneficial with a lower concentration of neutrophils and a higher concentration of CD4 T helper cells and salivary IgA. These biochemical indicator changes may be the critical mechanism for regular PA to lower the risk of hospitalization due to COVID-19.

As for the analysis of the confounding factors, results on between-subgroup heterogeneity showed significant heterogeneity in all subgroups adjusted for underlying disease. Thus, confounding factors of underlying disease significantly affected the association between PA and the risk of hospitalization. However, contrary to expectations, heterogeneity was observed in the within-subgroup analysis adjusted for age, sex, and BMI. This indicated that our data failed to demonstrate significant differences in the impact of PA on the risk of hospitalization for COVID-19 according to age, sex, and body weight. This may be because the overall heterogeneity of this meta-analysis was precisely concentrated in these subgroups. The other possible reason is that the effect of PA on reducing the risk of COVID-19 hospitalization is very stable among these different demographic characteristics.

The WHO global recommendation on the health benefits of PA states that adults have at least 150 minutes of moderate-intensity aerobic PA per week, at least 75 minutes of high-intensity aerobic PA per week, or a combination of moderate and high-intensity activities; this is equivalent to 10 met hour/week [1]. Our analyses also show that when the PA level is >10 met hours/week, the risk of hospitalization for COVID-19 is reduced, but the degree of reduction becomes significantly smaller. The practical significance of this conclusion is that PA or exercise within 10 met hours/week has the most apparent effect on reducing the risk of hospitalization for COVID-19, with an additional benefit for reducing the risk when PA level exceeds 10 met hours/week.

Increased hospitalization due to COVID-19 is a threat to health and a heavy disease burden on all aspects, such as individuals and the country. We found that increased physical activity significantly reduces the hospitalization risk of COVID-19, and physical activity should be a positive factor in decreasing the COVID-19 disease burden. However, the Global Burden of Diseases (GBD) 2019 ranked low physical activity 19th out of 20 risk factors in terms of disability-adjusted life years, down from 10th in the equivalent 2010 GBD publication [37,38]. Moreover, PA decreased in all age groups, independent of sex, due to the COVID-19 pandemic, according to a recent meta-analysis [39]. In the face of the global spread of COVID-19, we must regain the vital role of physical activity in reducing the burden of the disease. The conclusions of our study will undoubtedly have significant implications for public health.

Finally, our study is the first meta-analysis on the dose–response relationship between PA and the hospitalization risk of COVID-19. The results of this study are based on a large sample cohort study, case–control group study, and the advantages of a long period of follow-up investigation by cohort study; therefore, the results are relatively stable. However, this study may have limitations. First, the literature we included may be insufficient in the continuous dose–response analysis. One possible reason for this is that we set strict inclusion criteria. In addition, cohort studies require long-term follow-up surveys and extensive sample data. Therefore, few cohort studies have been conducted on related topics. Second, the methods of PA evaluation included in the literature of this study are subjective measurements, which may lead to inaccurate doses, and different measurement and observation methods of physical activities and hospitalized cases may have a significant impact on the research conclusion. In addition, exercise habit is based on the assumption that there is no change in the long-term follow-up; that is, the dose of PA is constant in the long-term observational study. This assumption may make the results inaccurate. Moreover, the results of some cohort studies included were not adjusted by the confounding factors such as sex, age, and other concomitant medical conditions, which should be paid attention to in future studies.

## 5. Conclusions

There was an inverse non-linear dose–response relationship between PA levels and the risk of COVID-19 hospitalization. An increase in the physical activity dose significantly reduced the hospitalization risk of COVID-19. The degree of risk reduction is weakened when PA is higher than 10 Met-h/week. Doses of the guideline-recommended minimum PA levels by the WTO may be required for more substantial reductions in the COVID-19 hospitalization risk. Future studies with different doses of PA or exercise interventions are needed to determine the optimum PA dose required for COVID-19 prevention.

## Data Availability

All data produced in the present work are contained in the manuscript

## Supporting information

S1 Checklist. PRISMA checklist of the meta-analysis.(DOC)

S1 Fig. Sensitivity analysis result. (TIF)

S2 Fig. Published bias funnel plot. (TIF)

## Acknowledgements

We would like to acknowledge the assistance of Chang Xu in helping us to develop an dose-response model with REMR approach.

## Author Contributions

**Conceptualization:** Dan Li, Songtao Lu.

**Data curation:** Dan Li, Shenzhen Jin, Songtao Lu.

**Formal analysis:** Shenzhen Jin, Songtao Lu.

**Methodology:** Dan Li, Shenzhen Jin.

**Software:** Dan Li, Shenzhen Jin. **Supervision:** Shenzhen Jin, Songtao Lu. **Visualization:** Dan Li, Shenzhen Jin.

**Writing – original draft:** Dan Li, Shenzhen Jin, Songtao Lu.

**Writing – review & editing:** Shenzhen Jin, Songtao Lu.

## Notes

### Competing Interest Statement

The authors have declared no competing interest.

### Funding Statement

This study did not receive any funding

### Author Declarations

Ethics committee/IRB of Full Institution name gave ethical approval for this work

